# A scoping review exploring the impact of digital systems on processes and outcomes in the care management of acute kidney injury and progress towards establishing learning healthcare systems

**DOI:** 10.1101/2021.02.23.21252060

**Authors:** Clair Chew, Helen Hogan, Yogini Jani

## Abstract

**Background:** Digital systems have long been used to improve the quality and safety of care when managing Acute Kidney Injury (AKI). The availability of digitised clinical data can also turn organisations and their networks into Learning Healthcare Systems (LHSs) if used across all levels of health and care. This review explores the impact of digital systems on AKI patient care to gauge progress towards establishing LHSs and to identify existing gaps in the research.

**Method:** Embase, PubMed, Medline, Cochrane, Scopus and Web of Science databases were searched. Studies of real-time or near real-time digital AKI management systems which reported process and outcome measures were included.

**Results:** Thematic analysis of 43 studies showed that most interventions used real-time serum creatinine (SCr) levels to trigger responses to enable risk prediction, early recognition of AKI or harm prevention by individual clinicians (micro level) or specialist teams (meso level). Interventions at system (macro level) were rare. There was limited evidence of change in outcomes.

**Conclusion:** Whilst the benefits of real time digital clinical data at micro level for AKI management have been evident for some time, their application at meso and macro levels is emergent therefore limiting progress towards establishing LHSs. Lack of progress is due to digital maturity, system design, human factors and policy levers. Future approaches need to harness the potential of interoperability and data analytic advances and include multiple stakeholder perspectives to overcome these factors.

## INTRODUCTION

The NHS was in the midst of a rapid phase of digital transformation before the Covid pandemic, which has patently further forced the pace of change.[1] The increasing availability of digitised clinical data has the potential to turn individual organisations and their networks into Learning Healthcare Systems (LHSs), systems that use information collected routinely as part of the care process to identify trends and variations and drive learning and quality improvement.[2] When this clinical information becomes near to or real time, it opens up the prospect not only of more detailed retrospective review of care but also the possibility of making more frequent and subtle adjustments across the system, to ensure quality is maintained as care proceeds.

The power of real-time clinical information to enable rapid adaptive responses to improve outcomes is already established at an individual patient level, for example, digitised Early Warning Scores reducing response time to deteriorating ward patients.[3] However for a LHS to be fully realised these data need to drive agile adaptation across different levels of the organisation and potentially the wider local health and social care system, facilitating changes that increase the chances of good outcomes for populations of patients whilst at the same time reducing risks of iatrogenic harm. Broadening “recognition and response” mechanisms from those focused on rapidly identifying and managing acute changes in individuals to real-time matching of acute illness burden to staff numbers and skill set on wards or converting hospital beds to higher care levels based on changes in demand is the next step towards building a LHS.[4] Limited progress in this direction has been reported, occurring mainly within individual organisations or healthcare systems rather than across the wider health and care system.[5]

Recent patient safety initiatives have prioritised detection and prevention of sudden deterioration, through focus on areas such as acute kidney injury management. Acute kidney injury (AKI) is a common complication found amongst acutely ill patients and has been associated with longer hospital stays, increased morbidity and mortality.[6] It can be a complication of an illness such as sepsis or a result of drugs or treatments the patient receives, especially where kidney function is already compromised by co-morbid illness.[7] There are no curative treatments but much can be done to limit kidney damage through institution of simple early interventions. This, in turn, avoids more complex interventions such as dialysis or renal replacement at a point where the kidneys can no longer be salvaged.

Diagnosis depends on a rising blood creatinine level or falling urine output. Laboratory values for creatinine can be easily digitised and the availability of electronic healthcare records (EHRs) have enabled the real-time/ near real-time reporting of values to clinicians. The NHS has recently introduced a standardised electronic reporting system for creatinine in an effort to decrease response times to treatment.[8] For EHRs that support clinical decision support systems (CDSS), computer physician order entry (CPOE) and electronic prescribing, alerts related to rising creatinine can be notified to the patient’s clinical team via the EHR providing real-time advice on an appropriate course of action and treatment choices.[9] Alternatively, such systems can send an alert to a pharmacist or renal rapid response team (RRT) to prompt action.[10,11] As well as promoting earlier diagnosis, some digital systems are predictive, identifying patients at risk and allowing closer monitoring or tailoring of treatment to avoid the condition developing.[12] Others play a part in harm-reduction by highlighting the potential dangers of certain drugs or doses to kidney function.

Given that digitisation of creatinine levels and real-time digital recognition and response systems for management of AKI have been available for over a decade, we used the literature to explore the extent to which such systems have impacted on patient care processes and outcomes across all levels of health and care systems (patient, organisation and population levels), to gauge progress towards the goal of establishing LHSs and to identify where current gaps in the research exist.

## METHODS

### Scoping Review

An initial scan of the literature on the use of real-time data for AKI management indicated a large variety of study approaches of varying methodology and rigour. A scoping review approach was selected to synthesise a metanarrative and identify themes based on the broad body of research in this field without exclusion based on study methods; a protocol was developed but not published.

### Search Strategy

Databases (Embase, PubMed, Medline, Cochrane, Scopus and Web of Science) were searched for papers published from inception to 31 January 2020 using free text keywords related to our review questions (Supplemental Material 1). Additional articles were identified through citation searches of relevant articles and reviews (Figure 1).

**Figure 1:**
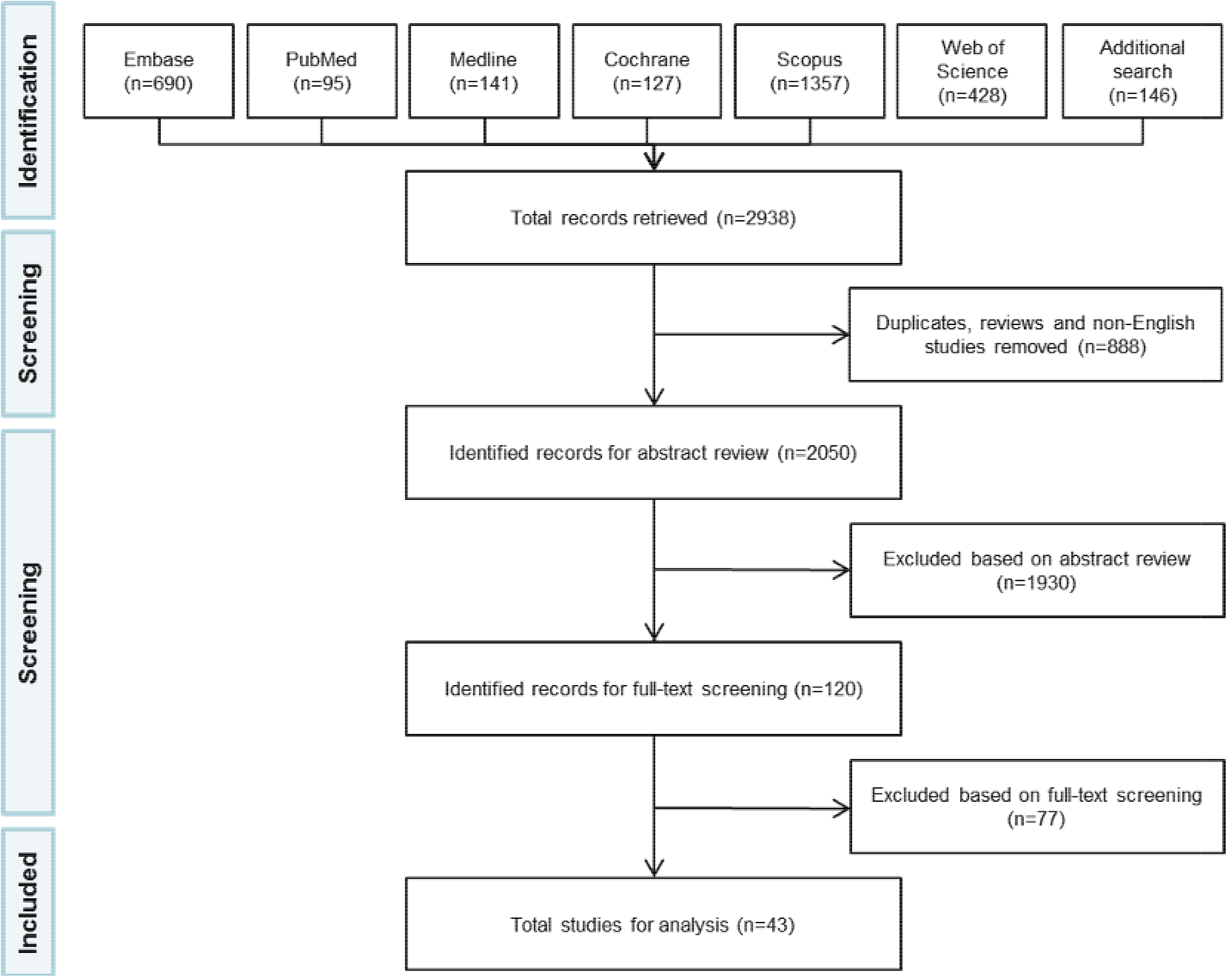
PRISMA flowchart of literature search.

### Study Selection

We included original research or case reports in the English language, conducted using any study design, in any setting, for any cohort of patients. We only included publications that reported process and/or outcome measures resulting from a real-time or near real-time healthcare professional response to data in the management of AKI e.g. interventions in medicines management in response to renal insufficiency. Reviews, narrative reports, observational or population health studies and publications which focused on model or alert development were excluded.

### Data Extraction

Our review objective was addressed through the following questions that formed a basis for thematic data extraction (Table 1).

**Table 1:** review questions mapped to themes used to analyse the studies.

| Review Question | Definition of concepts |
| --- | --- |
| At which level is the real-time data intended to generate action: what is the | **Micro: patient-level**<br>• clinical care and treatment at the patient level<br>**Meso: organisation/ specialty/service/unit management-level** |
| digital information designed to change? | For example,<br>• management of cohorts of renal patients by specialist e.g. pharmacist or renal specialist<br>• allocation of patients to a particular care pathway or ward<br>• staffing levels or skill mix<br>• resource distribution e.g. across diagnostic services or educational support or between harm management and risk assessment interventions<br>**Macro: population-level**<br>For example,<br>• targeting of interventions at particular populations e.g. primary or secondary care<br>• population management processes or the range of services that are available across the health and care system.<br>*NB. Some studies report on interventions where impact is intended at multiple levels. These were extracted to the higher level i.e. macro, meso then micro.* |
| What are the interventions and which staff are the targets? | **Afferent Arm (the monitored data item used to trigger a response)**<br>• Serum creatinine changes (SCr)<br>• Risk prediction score using composite values (on “entry” identify at risk of AKI before any treatments)<br>• Urine output<br>• Nephrotoxin exposure |
|  | **Timing (speed at which the digital data available to the responder)**<br>• Real time <1hour<br>• Near real time <24 hours |
|  | **Targeted group**<br>• Physician<br>• Nurse<br>• Pharmacist<br>• Two or more – multidisciplinary team<br>• Undefined (clinical team) |
| How integrated is the intervention into workflow? | **Efferent Arm (the alerting mechanism)**<br>• Interruptive within workflow<br>• Interruptive outside workflow<br>• Non-interruptive within workflow<br>• Non-interruptive outside workflow<br>• Undefined |
|  | **Level of digital maturity**<br>Level 1. Stand-alone afferent arm that requires human intervention for efferent mechanism such as by sending an email or text to raise an alert |
|  | Level 2. Integrated afferent and efferent arms in a single system with a specific focus such as e.g. pharmacy medicines management systems<br>Level 3. Integrated afferent and efferent arms that link alert data to wider response group across organisation or system but are not integrated into clinical workflow<br>Level 4. Integrated afferent and efferent arms that link alert data to wider response group across organisation or system and into clinical workflow<br>Level 5. Multi-organisation and cross-sectional (but otherwise same as 4) |
| Can use of real-time data improve processes of care and outcomes for AKI patients? | **Process Measure**<br>Measures of specific activity completed used in the study<br>**Outcome Measure**<br>Measures of clinical outcomes or proxies used in the study<br>**Findings**<br>Changes in process or outcome measures as a result of the intervention being studied |

SCr, serum creatinine.

- At which level is the real-time data intended to generate action: what is the digital information designed to change?
- What are the interventions and which staff are the targets?
- How integrated is the intervention into workflow?
- Can use of real-time data improve processes of care and outcomes for AKI patients?

## RESULTS

We identified 2050 unique articles (Figure 1). Following title and abstract screening using pre-specified criteria, 120 full text articles were reviewed, resulting in 43 studies (Supplemental Material 2) of interventions using real-time clinical information on AKI to drive service change and reported changes in either process or outcome measures (Table 2 and 3). The included studies were published between 1994 to 2020, with only seven publications before 2010.[13–19] The majority of studies were from the US and the UK, with 11 from other countries.[14,18,20–28] Most studies were conducted in hospitals with one in primary care,[29] and one involving community pharmacy services.[22] There were eight randomised controlled trials.[12,20,29–34] The other studies used a range of observational designs, with the majority being uncontrolled before and after studies.

**Table 2:** Thematic analysis of studies classifying the afferent arm, efferent arm, timing, targeted group, study type and level of digital maturity.

| Level | Purpose | Afferent arm | Efferent arm | Timing | Targeted group | Study type | Level of digital maturity |
| --- | --- | --- | --- | --- | --- | --- | --- |
| Micro | Risk prediction[12,46] | Risk prediction score [12,46] | Interruptive within workflow[12]<br>Non-interruptive within workflow[46] | Real time[12,46] | MDT[12,46] | Randomised controlled trial[12]<br>Controlled before and after[46] | 4[12,46] |
|  | Earlier diagnosis[14,20,24,25,30,31,40–45] | SCr[14,20,24,30,31,40–45]<br>SCr & urine output[25] | Interruptive within workflow[14,20,24,41–43]<br>Interruptive outside workflow[25,30,31,40]<br>Non-interruptive outside workflow[44,45] | Real time[14,20,25,30,31,41–43,45]<br>Near real time[24,40,44] | MDT[30,31,44]<br>Physician[14,24,25,40,41]<br>Undefined (clinical team)[20,42,43,45] | Randomised controlled trial[20,30,31]<br>Before and after[24,40,42,45]<br>Interrupted time-series[25,44]<br>Time-series[14]<br>Retrospective comparative study[43]<br>Observational descriptive study[41] | 1[40,44]<br>2[25,30,31,41,45]<br>3[24]<br>4[14,20,42,43] |
|  | Harm prevention[13,17,19,21,22,26,28,32,35–39] | Nephrotoxin exposure[19,39]<br>SCr[13,17,21,22,26,28,32,35–38] | Interruptive within workflow[17,19,22,26,28,32,37,39]<br>Interruptive outside workflow[35]<br>Non-interruptive within workflow[13,38,43]<br>Non-interruptive outside workflow[21] | Real time[13,17,19,22,26,28,32,35–39]<br>Near real time[21] | MDT[13,17,21,38]<br>Pharmacist[22]<br>Physician[19,26,28,32,35,37,39]<br>Undefined (clinical team)[36] | Randomised controlled trial[32]<br>Before and after[13,17,19,21,26,28,35,36,38,39]<br>Observational descriptive study[22,37] | 2[19,22,26,28,35,37,39]<br>3[13,21,38]<br>4[17,32,36] |
| Meso | Earlier diagnosis[11,49–52] | SCr [11,49–52] | Interruptive within workflow [49,50]<br>Interruptive outside workflow [11,52]<br>Non-interruptive outside workflow [51] | Real time[11,49,50,52]<br>Near real time[51] | MDT[11,49,52]<br>Physician[50,51] | Controlled before and after[11]<br>Before and after[49–51]<br>Qualitative interview study[52] | 2[11,50,51]<br>3[49]<br>4[52] |
|  | Harm prevention[10,15,16,23,27,33,47,48] | Nephrotoxin Exposure[10] SCr[15,16,23,27,33,47,48] | Interruptive within workflow[27]<br>Interruptive outside workflow[33,47]<br>Non-Interruptive within workflow[23,48]<br>Non-interruptive outside workflow[10,15,16] | Real time[33]<br>Near real time[10,15,16,23,27,47,48] | MDT[15,27,33]<br>Pharmacist[10,16,23,47,48] | Randomised controlled trial[33]<br>Controlled study[27]<br>Before and after[10,15,16,23,48]<br>Quality improvement[47] | 1[10]<br>2[15,16,23,27,47,48]<br>5[33] |
| Macro | Earlier diagnosis[29,53] | SCr[29,53] | Interruptive outside workflow[29,53] | Real time[53]<br>Near real time[29] | MDT[53]<br>Physician[29] | Quality improvement[53]<br>Randomised factorial design QI approach[29] | 5[29,53] |
|  | Harm prevention[34] | SCr[34] | Interruptive within workflow[34] | Real time[34] | Physician[34] | Randomised controlled trial[34] | 5[34] |

SCr, serum creatinine; MDT, multidisciplinary team; QI, quality improvement.

**Table 3:** Thematic analysis of studies highlighting the process measures and outcome measures used, and findings reported.

| Level | Purpose | Process Measures | Outcome Measures | Findings |
| --- | --- | --- | --- | --- |
| Micro | Risk prediction [12,46] | Changes in care management[46]<br>Frequency in monitoring or management[12]<br>Alert or recommendation generated/compliance[46] | AKI incidence[12,46]<br>AKI progression[46]<br>AKI severity[12]<br>Length of stay[12]<br>Mortality[12,46] | ↑ AKI documentation[46]<br>↑ Appropriate medication dosage[46]<br>↑ Proportion of SCr tests ordered[12]<br>↓ AKI incidence[46]<br>↓ Mortality[46] |
|  | Earlier diagnosis[14,20,24,25,31,40–45] | Detection[40,41]<br>Alert or recommendation generated/compliance[24]<br>Changes in care team or setting[20,45]<br>Changes in care management[14,31,40–44]<br>Appropriate care management[14,25]<br>Time to changes in care management[25,45] | AKI incidence[20,24,31,44]<br>AKI progression[25,31,42,43]<br>AKI recovery[24,25]<br>Duration of AKI[31]<br>Length of stay[25,31]<br>Mortality[24,25,30,31,42,43]<br>Change in SCr[30] | ↑ AKI documentation[31,44]<br>↑ AKI incidence[31,44]<br>↑ AKI recovery[24,25,31,42,43]<br>↑ Interventions[24,31,45]<br>↑ Rates of hospitalisation[45]<br>↓ Time to intervention[24,25]<br>↓ Length of stay[31]<br>↓ Mortality[42,43] |
|  | Harm prevention[13,17,19,21,22,26,28,32,35,37–39] | Detection[21]<br>Alert or recommendation compliance[17,32,37,39]<br>Changes in care management[17,21,22,26,32,35,38]<br>Appropriate care management[19,21,26,28,35,39]<br>Time to changes in management[13,39] | Rate of adverse drug events[36,37]<br>AKI progression[13]<br>Contrast induced AKI[26]<br>Length of stay[38]<br>Mortality[38] | ↑ Alert compliance[17,32]<br>↓ Alert compliance [37]<br>↑ Appropriate care management[17,21,22,32,35]<br>↓ Time to intervention[13]<br>↑ Care interventions[22]<br>↓ AKI progression[13]<br>↓ Length of stay[38]<br>↓ Mortality[38]<br>↓ Dialysis[38]<br>↑ Rate of potential adverse drug events[36]<br>↓ Rate of preventable adverse drug events[36] |
| Meso | Earlier diagnosis[11,49–52] | Detection[11]<br>Alert or recommendation generated/compliance[49]<br>Appropriate care management[49,52]<br>Changes in care team or setting[50,51] | AKI incidence[49]<br>AKI progression[49,50]<br>AKI recovery[11]<br>Cardiac arrest[50]<br>Change in renal function[50] | ↑ Alert compliance[49]<br>↓ Time to intervention[11,49]<br>↑ Recommendations[49]<br>↓ AKI incidence[49]<br>↓ AKI progression[49] |
|  |  | Changes in care management[50]<br>Time to changes in care management[11,49,51] | Early detection[52]<br>ICU admission[11]<br>Length of stay[51]<br>Length of stay cost[50]<br>Mortality[11,50,51]<br>Need for renal replacement therapy[11]<br>Peak SCr[51] | ↓ Time to AKI recognition[11]<br>↓ Possible cardiac arrests[50]<br>↓ Costs[50]<br>↑ Junior staff anxiety[52] |
|  | Harm prevention[10, 15,16,23,27,33,48] | Alert or recommendation generated/compliance[15,23,27]<br>Changes in care management[10]<br>Appropriateness of care management[15,16,23,33,48] | Adverse drug events[16]<br>AKI incidence[10,47]<br>Length of stay[16]<br>Cost of antibiotics[16]<br>Nephrotoxin exposure [10,47,48] | ↑ Appropriate care management[15,23,33,48]<br>↑ Care management interventions[10]<br>↑ Acceptance of recommendations[27]<br>↓ AKI intensity[10]<br>↓ Length of stay[16]<br>↓ Number of adverse drug effects[16] |
| Macro | Earlier diagnosis[29,53] | Detection[53]<br>Changes in care team or setting[29,53]<br>Changes in care management[53]<br>Alert or recommendation generated/compliance[53]<br>Patient given guidance[53] | AKI diagnosis[53]<br>Hospital Standardised Mortality Ratio[53]<br>Time to AKI response[29] | ↓ Mortality[29] |
|  | Harm prevention[34] | Appropriate care management[34] |  |  |

AKI, acute kidney injury; SCr, serum creatinine.

### Micro level

Thirty-two studies featured an intervention at the micro (individual patient) level. In 15 the main purpose of the intervention was harm prevention,[13,17,19,21,22,26,28,32,35–39] in 12 it was earlier diagnosis,[14,20,24,25,30,31,40–45] and in two, risk prediction.[12,46] Harm prevention interventions involved alerts to clinicians of the need to change nephrotoxic drugs (non-prescription, dose altering or drug suspension) based on a patient’s renal function. The main purpose of early diagnosis interventions was to alert individual clinicians of a patient’s deteriorating renal function to trigger an early review and appropriate intervention. Risk prediction interventions used algorithms to identify high risk individuals and institute individual management plans to prevent the development of AKI.

Interventions at this level were based on real-time data apart from four studies, which used near real-time data.[21,24,40,44] Three quarters of these interventions used interruptive alerts,[13,21,38,43–46] and in a third the alert was outside the clinicians’ workflow.[21,25,30,31,35,40,44,45] All early diagnosis alerts, apart from one (urine output [25]), were activated by changes in serum creatinine (SCr) levels. This was similar for harm prevention, with a minority of interventions using nephrotoxic drug exposure instead.[19,39] All the risk prediction interventions used algorithms to trigger alerts.[12,46]

In almost half the interventions where it was specified, the alert was targeted at a physician,[14,19,24–26,28,32,35,37,39–41] with a member of the multidisciplinary team (MDT) being the next most common target.[12,13,17,21,30,31,38,44,46] The digital maturity of the interventions clustered at level 2 (standalone databases not fully integrated into the EHR)[19,22,25,26,28,30,31,35,37,39,41,45] and level 4[12,14,17,20,32,36,42,43,46], two were at level 1[40,44] and four at level 3.[13,21,24,38]

### Meso level

Fourteen interventions were found at meso (management) level. Two thirds were harm prevention,[10,15,16,23,27,33,47,48] the others enabled earlier diagnosis.[11,49–52] Harm prevention interventions usually involved pharmacist surveillance of nephrotoxic medication across groups of patients at ward, specialty-unit or hospital level. Such surveillance led to patient intervention when kidney function was deteriorating and was often accompanied by feedback and education for clinical teams. Meso-level interventions aimed at early diagnosis were generally part of an approach to reducing the incidence and severity of AKI across a number of wards or the whole organisation. These interventions used the digital data in a variety of ways including to alert hospital-wide renal RRTs, to review patient management plans within ward-based safety huddles or to audit the timely implementation of AKI bundles (elements of protocolised AKI management plans). All but one of the interventions at meso-level used changes in levels of SCr to trigger an alert,[10] with two thirds based on near real-time activation,[10,15,16,23,27,47,48,51] and half being interruptive.[11,27,33,47,49,50,52] In five studies the alerts were presented within the clinical workflow.[23,27,48–50] The most popular recipient of the alerts was a pharmacist for harm prevention interventions and a member of the MDT for early diagnosis interventions. The digital maturity of interventions was low with the majority at level 2 and only three at level 3 or above.[33,49,52]

### Macro level

Just three studies had interventions that were designed to work at the macro (whole system) level.[29,34,53] Two focused on earlier diagnosis,[29,53] and one on harm prevention.[34] Two studies were based in the ambulatory care setting, one used alerts to notify primary care physicians of patients with AKI who needed review and the other identified contraindicated medication prescription in patients with compromised renal function. The third study described an organisation-wide quality improvement programme that included staff education, development of a care bundle and a renal RRT. All used changes in SCr level to trigger a response, all were interruptive, two thirds were real-time and targeted at physicians. These studies involved digital systems that spanned more than one organisation across the care system and therefore considered to have high digital maturity.

### Measures and outcomes

Study measures provide an implicit indication of the intervention goals. At the micro level, process measures for harm prevention interventions included adjustment of individual patient medication dose, completion of a medication review, and the time to medication adjustments or changes in monitoring regimes. Similar process measures were seen for early diagnosis and risk prediction interventions, focussing on changes in the recognition and recording of AKI, institution of appropriate individual patient management, and the timing of such actions or the timing between recognition of deterioration and escalation to higher acuity or specialist levels of care.

Process measures at the meso level were similar to those seen for micro harm prevention interventions, with the addition of measures reflecting the degree of acceptance of pharmacist recommendations by physicians. Meso-level interventions that focused on early diagnosis used process measures such as time to AKI recognition, the percentage of changes made across the care pathways of interest, number of activations of renal RRTs and the time between team activation and patient intervention. AKI detection rate and clinician engagement with renal RRTs were process measures for early diagnosis interventions at the macro level. For harm prevention interventions, the proportion of inappropriately prescribed nephrotoxic drugs was measured.

Outcome measures were similar across all system levels and included AKI rates, AKI severity, rates of recovery, progression, initiation of renal replacement therapy, admissions to higher acuity or specialist care, length of stay and mortality. For harm prevention interventions this was supplemented with proportions of adverse events.

The impact of the interventions was mixed. Amongst micro-level interventions over half of early diagnosis interventions showed positive changes in outcomes.[24,25,30,42–44] Only one study was a randomised control trial (RCT)[31] and this showed a reduced length of stay. One third of harm prevention studies at this level found improvements in outcomes,[13,36,38,46] none of which were RCTs. Two out of three risk identification studies had a positive impact on outcomes. At the meso level there were no high-quality studies. One fifth of harm prevention[10,16] and two-fifths of early diagnosis[49,50] interventions had the desired impact. At the macro level, one RCT found a reduction in mortality following an ambulatory care intervention to increase the recognition of AKI.[29] Across harm prevention interventions at all levels there was evidence of a positive change in the most common process measures (reduced prescription of nephrotoxic medication and more appropriate dosing) in 42% of studies.[13,15,48,17,18,21–23,32,33,46]. Fewer earlier diagnosis intervention studies (29%) showed positive findings for the most common process measures (time to recognition and response to AKI and institution of more elements of appropriate management).[11,24,25,31,45,49]

## DISCUSSION

Given the longstanding availability of AKI digital information we used this condition to examine how digital clinical systems were maturing towards LHS. Our findings show that whilst such systems have had a positive effect for over 30 years at micro levels, their application at macro levels is emergent. Most interventions used SCr levels to trigger alerts or algorithms in real or near real time to enable risk prediction, early recognition of AKI or harm prevention by individual clinicians or specialist teams such as pharmacists and renal RRTs. Evaluations using process measures indicate apparent gains in harm reduction through avoidance of nephrotoxic medications or doses, or earlier prediction of the risk of deterioration. Evidence for improved outcomes is limited, with change more often seen in proximal outcomes such as length of stay in the lower quality studies and a few studies reporting reduction in mortality.[29,38,42,43,46,53] Much remains to be understood about the longevity and sustainability of the interventions, but there are signals that this may be feasible within integrated health systems.[53]

The limited evidence on interventions and positive outcomes at the meso and macro level may be explained by several factors. Many digital systems have evolved from clinician interest in better management of individual patients and recognition that the ‘right’ data needs to be presented in an appropriate format, in a timely manner at the appropriate point in the workflow. Thus, the majority of reported interventions were targeted at individual clinicians or specialist teams, using changes in SCr as the trigger. Expansion of the use of real-time digital clinical information to improve quality of care at meso and macro levels will also require the increasing digital maturity of systems. With the transition from standalone to integrated EHR within and across health systems more data will be available not just to clinicians at the point of care, but also the wider multidisciplinary team (MDT) as well as organisation and system managers.

However, data alone is insufficient for changing or influencing behaviours. Recognising and understanding the role of human factors in EHR design and utilisation is important to ensure maximum benefit of real-time data at relatively neglected meso and macro levels. Furthermore, challenges of generating actionable data include considerations of how the data are conveyed to enable a real-time response from the most appropriate persons. In the evidence reviewed, many systems relied on interruptive alerts or alerts that were outside the clinicians’ workflow. Other reviews have highlighted that success of alerts and accompanying clinical decision support systems to change user behaviours is dependent on workflow integration, level of intrusiveness and presence of multiple competing alerts, with alert fatigue cited as the most frequent reason for ineffectiveness.[54,55]

Successful transition from data utilisation to data driven healthcare has implications for technical factors (system design), human factors (behavioural impact) and resources (individuals, infrastructure), and requires a supportive, adaptive policy environment.[56] Advances in technical factors through EHR systems within organisations are becoming established but need to progress towards integration and interoperability across organisations and with other systems, such as management databases for staffing. A range of disciplines need to be involved in further developments, including clinicians, human factors experts, behavioural scientists, technology experts and data scientists. Developing the analytics capability and digital literacy of clinical and administrative staff is fundamental for successful LHSs, to develop mechanisms to monitor the impact of the use of information and to enable continuous tailoring (to different contexts and staff compositions), especially in the light of changing contexts and the need to respond to user feedback.

The recent experience of the Covid-19 pandemic illustrates that under these unusual conditions adaptive and enabling policies, with the rapid development, deployment, and innovative use of digital systems can enable continuity of healthcare delivery across acute and primary care sectors. Other examples of data-driven enabling policies at macro level such as the UK value-based commissioning,[57] or ‘getting it right first time’ programmes,[58] demonstrate the feasibility of using routinely collected clinical data at system level to determine care outcomes or to better understand the causes of their variation, signalling what might be possible within an effective digital LHS.

From a research perspective, evidence is needed from studies that go beyond immediate care settings expanding measurement to indicators of system dependent health outcomes such as hospital avoidance, reduced length of stay and access to healthcare services. Well chosen patient-centred process and outcome indicators from across the system will provide feedback in real time to steer individual patient care, as well as provide information that may be available later for reflective and responsive learning at population level, from small groups of patients up to larger populations. This requires a different real-time focus on the same data, promoting reactive behaviour at the micro level whilst also providing insight into variations that may be addressed at meso and macro levels through adaptive changes in service delivery and resource (re)distribution.

### Strengths and Limitations

Our scoping literature review format combining clearly defined key concepts and a systematic approach enabled exploration and synthesis of a complex and heterogeneous area and the capture of most relevant and appropriate articles. However, there may be examples of the use and impact of real-time data at meso and macro level not published in academic literature, as developments at these levels are relatively immature. Moreover, we may have misclassified some intervention across micro, meso and macro levels as the interventions were not always well described. It was also not our intention to formally assess the quality of included papers given that we were as interested in which dimensions of intervention process or outcomes were chosen for measurement as we were in the impact of the intervention. In the majority of cases, drawing conclusions about the latter was difficult given the limitations of study designs used.

## CONCLUSIONS

Digital transformation, use of data in real time and LHSs are cornerstones for achieving the triple aim to improve population health, quality of care and cost control.[59–61] Wider approaches are now required to build on the initial impact seen at individual patient level in order to gain benefits across the system, particularly in service delivery and resource distribution. This will require a coordinated effort across developments in technical, human factor and policy arenas with adequate resourcing. The lessons learned from deployment of digital systems to enable the coordination of resources across primary and secondary care during the Covid-19 pandemic should act as a powerful catalyst.

## Data Availability

All data relevant to the study are included in the article or uploaded as supplementary information

## ACKNOWLEDGEMENTS

At the time of completing this work, Yogini Jani and Helen Hogan were Health Foundation Improvement Science Fellows. The Health Foundation is an independent charity committed to bringing about better health and health care for people in the UK.

