## Supplemental Material for "A scoping review exploring the impact of digital systems on processes and outcomes in the care management of acute kidney injury and progress towards establishing learning healthcare systems"

### Supplemental Material 1: Search terms

Databases (Embase, PubMed, Medline, Cochrane, Scopus and Web of Science) were searched for papers published from inception to 31 January 2020 using free text keywords related to our review questions. Additional articles were identified through citation searches of relevant articles and reviews.

Search terms used were:

(Acute Kidney Injury or Acute Renal Failure or Renal or AKI)

and

(Decision Support or health information exchange or hospital information system or EHR or EPR or electronic or computer* or CPOE or Surveillance or Monitoring or Detection or Management or Prevention or Prescribing or Treatment or Alert* or predictive analy* or  predictive model* or machine learning or care process models or resource utilisation or clinical workflow or referral tracking or hospital service or care models or intervention).

### Supplemental Material 2: Summary of the 43 studies included in the scoping review

**Table S1: Summary of 43 studies included in the scoping review**

| **Surname** *et al* Country^citation number | **Population & Setting** | **Intervention** |
| --- | --- | --- |
| Aiyegbusi et al UK (Scotland) (2018)[1] | All primary care in NHS Tayside region | AKI identified in primary care (PC-AKI) through AKI e-alerts |
| Al-Jaghbeer et al USA (2018)[2] | 14 hospitals in a health care system | Clinical decision support system in hospital |
| Awdishu et al USA (2016)[3] | Tertiary healthcare hospital and ambulatory care system | Clinical decision support tool developed for 20 nephrotoxic medications |
| Bhardwaja et al USA (2011)[4] | Single large integrated health care delivery system (Kaiser Permanente Colorado) | Use of pharmacy alert system to reduce medication errors in renal insufficiency |
| Chandrasekar et al UK (2017)[5] | Acute hospital admissions | Whole system quality improvement approach |
| Chertow et al USA (2001)[6] | Urban tertiary care teaching hospital | Computerised decision support for prescribing in patients with renal insufficiency |
| Cho et al Republic of Korea (2012)[7] | Teaching hospital | Computer alert for risk of contrast-induced AKI and recommendation for prophylaxis |
| Choi et al Republic of Korea (2019)[8] | Hospital patients with eGFR less than 50 | Designated pharmacist in addition to computerised alerts |
| Colpaert et al Belgium (2012)[9] | Tertiary Hospital | Introduction of real-time electronic alert system to improve management and severity of AKI |
| Connell et al UK (2019)[10] | A large hospital | A digitally enabled care pathway comprising automated AKI detection, mobile clinician notification, in-app triage, and a protocolised specialist clinical response. |
| Connell et al UK (2019)[11] | ED departments in single tertiary hospital (intervention) vs single district hospital (control) | Multicomponent intervention (alert system, AKI response team and care protocol) to improve the outcomes from AKI |
| Connell et al UK (2019)[12] | Tertiary care hospital | Mobile results viewing in a digitally enabled care pathway |
| Desmedt et al Belgium (2018)[13] | Academic hospital (non-ED or ICU patients) | Computerised decision support for dosing adjustments for 85 drugs |
| Díaz et al Spain (2013)[14] | Teaching hospital | A system for drug dosage adjustment integrated into the hospital computer provider order entry system |
| Evans et al USA (1999)[15] | Tertiary care centre | Computer-assisted antibiotic dose monitor |
| Galanter et al USA (2005)[16] | Single teaching hospital | Automated alerts designed to reduce the use of contraindicated drugs in patients with renal insufficiency |
| Goldstein et al USA (2013)[17] | Single quaternary paediatric hospital | Pharmacist screen for nephrotoxic load and recommendations for serum creatinine testing made |
| Goldstein et al USA (2016)[18] | Children noncritical care unit | Pharmacist recommended monitoring and dosing after electronic trigger |
| Heringa et al Netherland (2017)[19] | Community pharmacies | CDSS with optional point of care testing |
| Hodgson et al UK (2018)[20] | 2 non-specialist hospitals | Electronic clinical prediction rule combined with an AKI e-alert |
| Kolhe et al UK (2015)[21] | Tertiary care centre | A care bundle with interruptive alert |
| Kolhe et al UK (2016)[22] | Single teaching hospital | AKI care bundle with interruptive alert |
| Kothari et al USA (2018)[23] | 8 New York hospitals | Daily laboratory alerting of patients at risk for AKI |
| Leung et al USA (2013)[24] | 5 Community Hospitals | Comparison of different intensities of clinical decision support within EHR computer order physician entry |
| Matsumura et al Japan (2009)[25] | Single hospital | Development of EHR e-alert system for evaluating renal function and checking doses of medication according to the patient's renal function |
| McCoy et al USA (2010)[26] | Academic tertiary care hospital | Computerised order entry alerts: passive alert for increasing creatinine and interruptive alert for medication adjustment |
| Nash et al USA (2005)[27] | Teaching hospital | An automated system to complement an existing computerized order entry system by detecting the administration of excessive doses of medication |
| Park et al Korea (2018)[28] | Tertiary teaching hospital | AKI alert system that provides option for automated consultation requests to the nephrology division |
| Porter et al UK (2014)[29] | Teaching hospital | Real-time alert to detect AKI |
| Ralph et al USA (2014)[30] | Tertiary hospital | Pharmacist-run CDSS alert based on early serum creatinine |
| Rind et al USA (1994)[31] | Teaching hospital | Computer-based alerts for hospitalised patients |
| Roberts et al Australia (2010)[32] | Teaching hospital | CDSS in an environment independent of computerised provider order entry introduced to prescribers via academic detailing |
| Selby et al UK (2019)[33] | 5 hospitals | Multi-faceted intervention programme (AKI e-alerts, an AKI care bundle, and an education program) to improve outcomes associated with AKI |
| Sellier et al France (2009)[34] | 2 departments in a teaching hospital | Alert at time of ordering medication in computerised provider order entry system to decrease inappropriate prescriptions |
| Sykes et al UK (2018)[35] | Single teaching hospital | Whole system approach quality improvement to reduce AKI and its impact (e-learning package, AKI bundle, enhanced pharmacy medicines reconciliation, QI nurses, safety huddles, supporting literature, champions) |
| Thomas et al UK (2015)[36] | 2 acute hospitals and a community service | AKI outreach service |
| Tollitt et al UK (2018)[37] | 46 primary care practices | AKI e-alert and AKI educational outreach sessions |
| Van Driest et al USA (2020)[38] | Teaching hospital | Implementation of AKI risk alerts to promote increased uptake of serum creatinine screening of patients |
| Vogel et al USA (2016)[39] | Integrated health care delivery system (Kaiser Permanente Colorado) | Pharmacy and physician facing e-alert system linked to prescribing |
| West Midlands Acute Medicine Collaborative et al UK (2019)[40] | Acute medical units in 14 hospital sites | UK National Health Service AKI e-alert system |
| Wilson et al USA (2015)[41] | Tertiary Hospital | Use of automated e-alert to reduce severity of AKI injury and improve outcomes |
| Wong et al USA (2017)[42] | Urban tertiary care hospital ICU | Computerised decision support including to support safe drug use in renal insufficiency |
| Wu et al China (2018)[43] | Hospital ICUs and high-risk cardiovascular wards | AKI e-alert on high-risk wards |
| AKI, acute kidney injury; PC-AKI, primary care acute kidney injury; GFR, glomerular filtration rate; ED, emergency department; ICU, intensive care unit; EHR, electronic health system; CDSS, clinical decision support system. | | |
